# Cross-ancestry evaluation of idiopathic pulmonary fibrosis genetic risk variants

**DOI:** 10.64898/2026.04.17.26349970

**Authors:** Ritah Nabunje, Beatriz Guillen-Guio, Tamara Hernandez-Beeftink, Ebrima Joof, Olivia C Leavy, International IPF Genetics Consortium, Toby M Maher, Phil Molyneux, Justin M Oldham, Imre Noth, Amaia Urrutia, Myriam Aburto, Carlos Flores, R Gisli Jenkins, Louise V Wain, Richard J Allen

**Affiliations:** Division of Public Health and Epidemiology, University of Leicester, Leicester, UK; University Hospitals of Leicester NHS Trust, Leicester, UK; Centre for Fibrosis Research, University of Leicester, Leicester, UK; Centro de Investigación Biomédica en Red de Enfermedades Respiratorias (CIBERES), Instituto de Salud Carlos III, Madrid, Spain; NIHR Imperial Biomedical Research Unit, National Heart and Lung Institute, Imperial College London, London, UK; Keck Medicine of USC, University of Southern California, Los Angeles, California, USA; University of Michigan, Michigan, USA; University of Virginia, Virginia, USA; Servicio de Neumología, Hospital Universitario de Cruces, Barakaldo, Vizcaya, Spain; Departamento de Medicina, Universidad del País Vasco, Spain; Servicio de Neumología, Biobizkaia, Hospital Universitario de Galdakao-Usansolo, Usansolo, Vizcaya, Spain; Departamento de Medicina, Universidad del País Vasco, Spain; Genomics Division, Instituto Tecnologico y de Energias Renovables, Santa Cruz de Tenerife, Spain; Research Unit, Hospital Universitario Nuestra Señora de Candelaria, Instituto de Investigación Sanitaria de Canarias, Santa Cruz de Tenerife, Spain; Facultad de Ciencias de la Salud, Universidad Fernando Pessoa Canarias, Las Palmas de Gran Canaria, Spain

## Abstract

Genome-wide association studies of idiopathic pulmonary fibrosis (IPF) have identified 35 common genetic risk loci associated with IPF susceptibility. In this study, we evaluated the effects of the reported variants in clinically curated non-European individuals. Despite limited sample sizes, we observed partial replication, limited transferability of some variants and evidence of ancestry-specific effects. The *MUC5B* promoter variant rs35705950 emerged as the dominant and most consistent signal across ancestries. Our findings highlight the need for larger, well-characterised studies in understudied populations to support robust discovery and translation.

## Introduction

Previous genome-wide association studies (GWAS) of idiopathic pulmonary fibrosis (IPF) have identified 35 common genetic risk loci associated with IPF susceptibility (1, 2). However, these studies have been conducted in cohorts predominantly of European genetic ancestry. A recent multi-ancestry GWAS for IPF has been conducted by Partanen et al 2022 (1), however this used cases defined using electronic health records which often lead to imprecise effect estimates for IPF (3). We therefore sought to evaluate the effects of known genetic risk variants in well-defined non-European individuals.

## Methods

Non-European cases were selected from IPF cohorts previously described (2). We also used cases from an isolated European population from the Basque Country in Northern Spain, genotyped on the Axiom Spain Biobank Array. All cases were curated by clinicians following established diagnosis guidelines (4).

A random forest classifier was trained using the first ten principal components (PCs) derived from the 1000 Genomes Project (1000GP) (5) super populations (African [AFR], Admixed American [AMR], East Asian [EAS], European [EUR] or South Asian [SAS]), using 10-fold cross validation and an 80:20 training-testing split. The classifier was used to estimate the posterior probability of the study individuals’ membership in each ancestry group. Individuals were assigned to an ancestry group if the predicted probability exceeded 50% for that group.

Within each non-European ancestry group, cases were matched to ten controls of the same ancestry from UK Biobank (6) based on the first ten principal components. Basque Country IPF cases were matched to controls in the National DNA Bank of Spain (7). Samples were screened for relatedness, and one individual from each 2^nd^-degree related pair (kinship coefficient ≥0.0884) was removed. Genotype imputation was performed using the TOPMed R3 reference panel (8) on the Michigan Imputation Server (9) using variants common to all cases and controls.

A set of 35 independent genome-wide significant variants were selected from IPF susceptibility studies (1, 2). The allele frequencies (AFs) for these variants were calculated in each of the IPF case-control ancestry groups and compared to AFs in 1000GP.

Logistic regression was performed to test the association of each variant with IPF in each ancestry group using PLINK2 (10), adjusting for population structure using the first ten PCs. Variants with a low imputation score (r^2^<0.6) were excluded. To assess heterogeneity in genetic effects across populations, trans-ethnic meta-regression was performed using MR-MEGA for variants that were present in at least four ancestry groups. Statistical significance was assessed using a Bonferroni corrected p-value=1.4×10^−3^ to account for multiple testing.

The cumulative effect of the 35 variants was assessed using a polygenic risk score (PRS) constructed and tested for association with IPF separately within each ancestry group using PRSice-2 (11). The contribution of the PRS to IPF status was quantified using Nagelkerke’s pseudo-R^2^. Given its large effect size, PRS were also calculated excluding the *MUC5B* variant.

## Results

We included 41 AFR, 61 SAS, 21 EAS and 92 AMR ancestry individuals with a clinical diagnosis of IPF. From the Basque cohort, 98 EUR ancestry samples were included. Due to relatedness, 14 individuals were removed. Across most ancestry groups, cases were more likely to be male, except for AFR (**Table 1**).

**Table 1:** Sample characteristics and number of analysable IPF risk variants by genetic ancestry. Summary of the number of IPF cases and controls, mean age at assessment, and sex distribution presented as the number and percentage of male participants across ancestry groups. The number of variants included in downstream analyses reflects exclusion of monomorphic variants (AF = 0) and variants with low imputation quality (r^2^ < 0.6).

|  | AMR | SAS | AFR | EAS | EUR |
| --- | --- | --- | --- | --- | --- |
| **Sample size** |  |  |  |  |  |
| Cases | 90 | 60 | 41 | 21 | 98 |
| Controls | 915 | 608 | 409 | 208 | 979 |
| **Mean age** |  |  |  |  |  |
| Cases | 66.2 | 64.8 | 58.1 | 75.1 | 69.8 |
| Controls | 52.4 | 54.2 | 51.9 | 51.7 | 57.5 |
| **Male sex, N (%)** |  |  |  |  |  |
| Cases | 64<br>(71.1%) | 50<br>(83.3%) | 17<br>(41.4%) | 14<br>(66.7%) | 81<br>(82.7%) |
| Controls | 489<br>(53.4%) | 490<br>(80.6%) | 186<br>(45.5%) | 137<br>(65.9%) | 819<br>(83.7%) |
| **Number of variants included in the analyses** | 34 | 33 | 34 | 24 | 34 |

The association of each of the 35 genetic variants previously associated with IPF was calculated (**Figure 1**). Due to differences in allele frequency and imputation quality, the number of variants analysed for each population varied (**Table 1**). Variants rs539683219 (*PSKH1*) and rs115610405 (*RTEL1*) were monomorphic in one or more populations.

**Figure 1:**
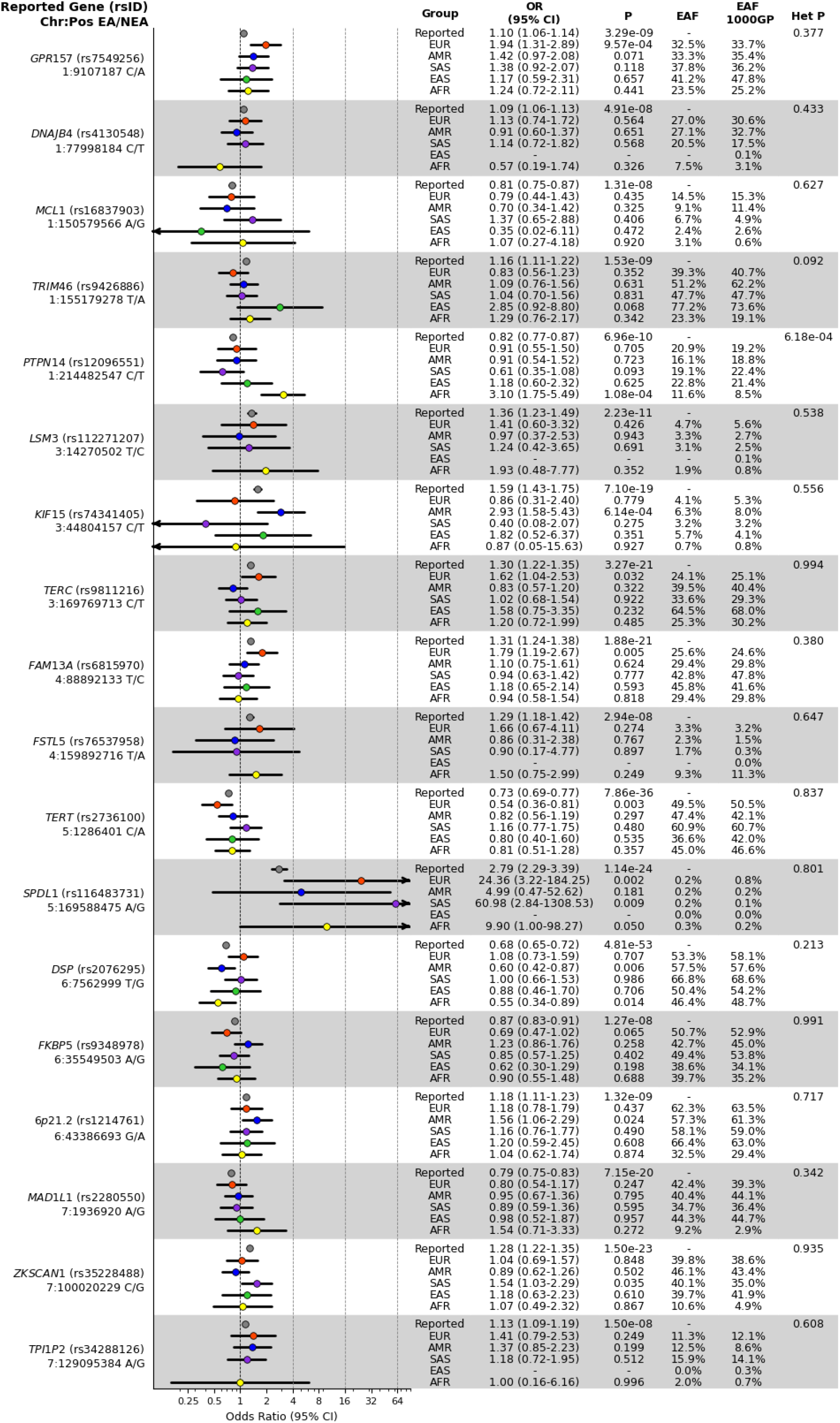
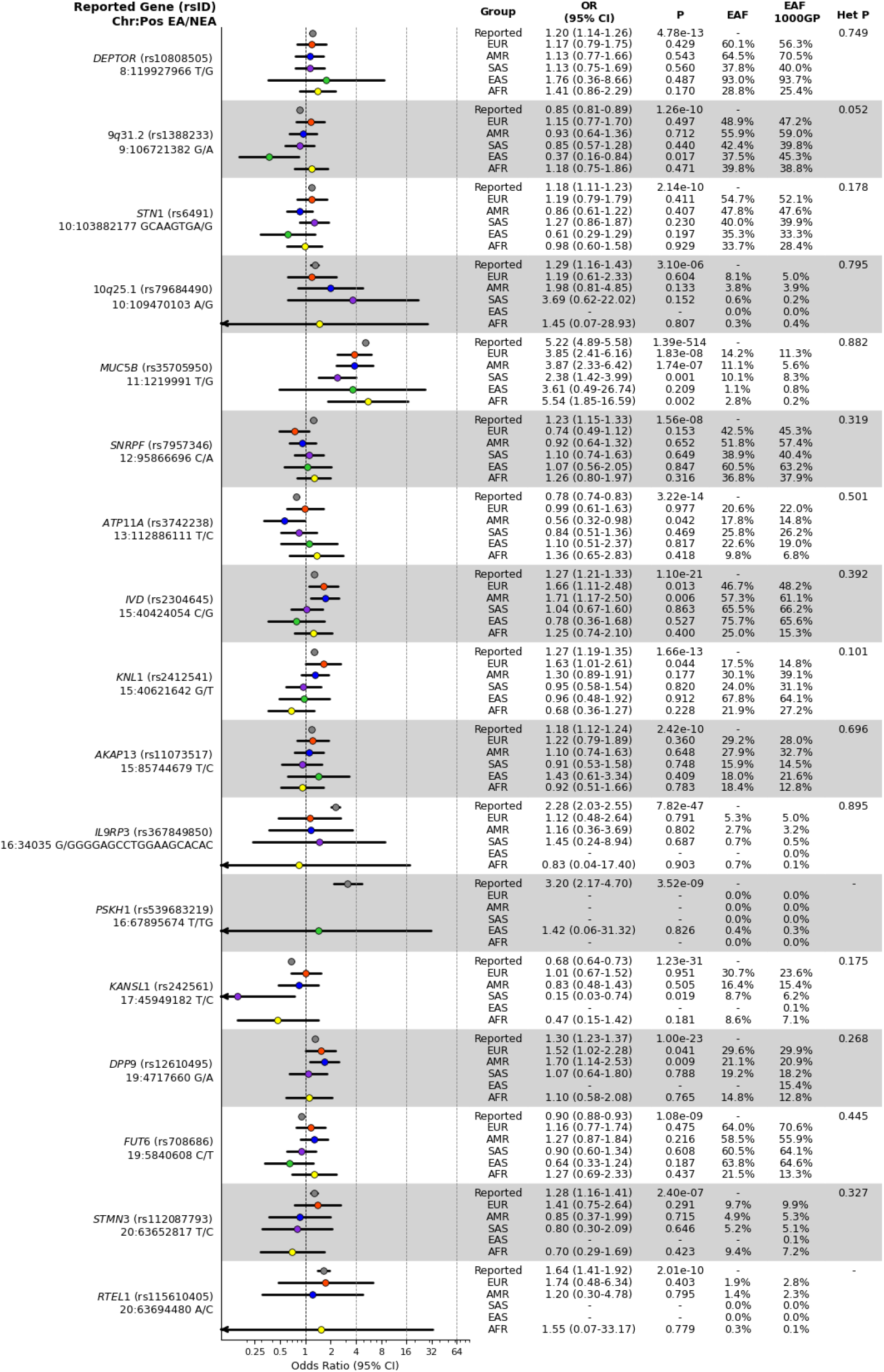
Single-variant association effect estimates for IPF risk variants across ancestry groups. Effect allele (EA), non-effect allele (NEA), odds ratio (OR) and 95% confidence intervals (CI) for each variant and population, alongside the corresponding p-values as well as the effect allele frequency (EAF) in the study cohorts and 1000GP populations, and the p-values of ancestry-correlated heterogeneity (Het P). Effect size estimates are coloured by ancestry group with the previously reported effect sizes (“Reported”) shown in grey. For all variants, reported effect size is taken from the Chin et al GWAS of clinically-curated European IPF cases apart from GPR157 (rs7549256), DNAJB4 (rs4130548), FSTL5 (rs76537958), TPI1P2 (rs34288126), PSKH1 (rs539683219) and FUT6 (rs708686) for which reported effect size is taken from the Partanen et al. (2022) multi-ancestry GWAS meta-analysis.

In the EAS cohort, 11 of 35 variants were excluded: seven with imputation quality below the r^2^ < 0.6 threshold and four that were monomorphic. Consistent with 1000GP, rs79684490 (*10q25.1*) and rs116483731 (*SPDL1*) were monomorphic in EAS. The rs34288126 (*TPI1P2*) variant was absent in our EAS cohort but is present in 1000GP EAS, albeit at very low frequency (AF=0.26%).

The *SPDL1* missense variant rs116483731 exhibited the strongest effects across the populations (**Figure 1**) with point estimates exceeding those previously reported. The *MUC5B* promoter variant rs35705950, the strongest genetic risk factor for IPF in Europeans, showed consistent effects across ancestry groups (**Figure 1**) despite substantial differences in AF.

The *PSKH1* variant (rs539683219), previously associated with IPF in EAS by Partanen et al., (2022), was polymorphic only in EAS. However, it was not significantly associated with IPF risk in our study and effect estimates were imprecise.

The only variant with significant heterogeneity of effect sizes across populations was the *PTPN14* variant rs12096551 (P=6.2×10^−4^) (**Figure 1**). The C-allele previously associated with decreased risk of IPF in European cases was associated with increased risk of IPF in the AFR (Odds Ratio=3.10, 95% Confidence Interval=1.75-5.49, P=1.08×10^−4^).

The PRS was associated with IPF risk in all ancestry groups with the largest Nagelkerke’s R^2^ observed in EUR (R^2^=0.09, P=1.2×10^−10^) and AMR (R^2^=0.08, P=6.9×10^−9^) and smaller values in SAS (R^2^=0.05, P=7.1×10^−5^), EAS (R^2^=0.05, P=0.021) and AFR (R^2^=0.03, P=0.006). Exclusion of the *MUC5B* variant reduced the variance explained by the PRS in all populations. The PRS excluding the *MUC5B* variant remained associated with IPF risk in EUR (R^2^=0.03, P=1.9×10^−5^), AMR (R^2^=0.03, P=4.9×10^−5^), SAS (R^2^=0.02, P=0.025) and EAS (R^2^=0.04, P=0.038) but showed no evidence of association in AFR (R^2^=0.01, P=0.133).

## Discussion

Here, we report effect estimates of previously reported IPF genetic risk variants across diverse clinically curated IPF cases drawn from cohorts, registries and clinical trials. Several variants identified in European populations were monomorphic in other populations underscoring limited transferability. The *MUC5B* promoter variant rs35705950 emerged as a dominant contributor to IPF genetic risk across populations. Evidence of ancestry-correlated heterogeneity for the *PTPN14* variant suggests possibility of population-specific genetic effects. The EAS-specific *PSKH1* variant (rs539683219) reported by Partanen et al. (2022) was not significant in this study most likely reflecting small EAS sample size.

The PRS combining variant effects was associated with IPF risk across ancestry groups with differing proportions of variance explained. Exclusion of the *MUC5B* variant substantially reduced the association in EUR, AMR, and SAS, negated the association in AFR but had minimal impact in EAS. The weaker or non-significant PRS association in SAS and AFR may reflect reduced portability of reported effect sizes.

Several limitations should be considered. Firstly, sample sizes were modest, limiting statistical power and precision of estimates. Secondly, analyses were restricted to previously reported variants, biasing results toward discoveries in predominantly European populations. Finally, variation in imputation quality reduced analysable variants across ancestries and PRS construction did not account for ancestry-specific effect sizes or LD patterns.

These findings provide a foundation for future work which will require larger, well-curated cohorts from diverse populations to enable robust genetic discovery and assessment of generalisability of IPF genetic risk factors across populations.

## Data Availability

All data produced in the present study are available upon reasonable request to the authors.

## Funding and acknowledgements

This study was funded by a Wellcome Trust PhD studentship for RN as part of the Wellcome Trust Genetic Epidemiology and Public Health Genomics Doctoral Training Programme (218505/Z/19/Z). RJA is supported by UK Research and Innovation grant UKRI1481. LVW held a GlaxoSmithKline / Asthma + Lung UK Chair in Respiratory Research (C17-1). BGG is supported by Wellcome Trust grant 221680/Z/20/Z. JMO reports National Institutes of Health (NIH) National Heart, Lung, and Blood Institute grants R56HL158935 and K23HL138190. CF is supported by the Instituto de Salud Carlos III (PI20/00876, PI23/00980, CB06/06/1088, and PMP22/00083), co-financed by the European Regional Development Funds “A way of making Europe” from the EU, and by an agreement with Instituto Tecnologico y de Energias Renovables (ITER) to strengthen scientific and technological education, training, research, development and innovation in genomics, epidemiological surveillance based on massive sequencing, personalized medicine, and biotechnology (OA23/043). This work was partially supported by the National Institute for Health Research (NIHR) Leicester Biomedical Research Centre (NIHR203327). The views expressed are those of the author(s) and not necessarily those of the National Health Service, the NIHR, or the Department of Health and Social Care. This research used the ALICE and SPECTRE High Performance Computing Facility at the University of Leicester.

